# Automatic Contact Tracing for Outbreak Detection Using Hospital Electronic Medical Record Data

**DOI:** 10.1101/2020.09.08.20190876

**Authors:** Michael DeWitt

## Abstract

Contact tracing is a well-known tool for public health professionals to trace and isolate contacts of known infectious persons. During a pandemic contact tracing is critical to ending an outbreak, but the volume of cases makes tracing difficult without adequate staffing tools. Hospitals equipped with electronic medical records can utilize these databases to automatically link cases into possible transmission chains and surface potential new outbreaks. While this automatic contact tracing does not have the richness of contact tracing interviews, it does provide a way for health systems to highlight potential super-spreader events and support their local health departments. Additionally, these data provide insight into how a given infection is spreading locally. These insights can be used to inform policy at the local level.

## 1 Introduction

Contact tracing is an effective way to trace the origin of cases and is a well known tool for public health. Through contact tracing, infected individuals are isolated and a history of who an infected individual had close contact with is established. Individuals with close contact are then interviewed for symptoms and isolated themselves in order to arrest additional transmission of an infection. Effective use of contact tracing can drastically reduce the duration of infection (Eames and Keeling 2003). However, contact tracing typically relies on small staffs of local public health offices. During the SARS-CoV-2 outbreak, these public health offices were and continue to be saturated with cases. With each non-isolated positive case making roughly 10 (Jarvis et al. 2020) contacts per day^1^ and a serial interval of 4-5 days (Nishiura, Linton, and Akhmetzhanov 2020; Bi et al. 2020), this means that for each positive case, somewhere between 20-40 unique contacts need to be tracked per new case, assuming that cases are detected at the first sign of symptoms. In the middle of an outbreak the workload is daunting. Effective contact tracing is a part of the United States Center for Disease Control’s (CDC) guide (Disease Control and Prevention 2020) to relaxing restrictions on social distancing.

Automated contact tracing is not a new idea, however, Electronic Medical Records (EMR) have not been adequately explored as an option for rapid contact tracing during an outbreak. Previous research found that use of EMRs could lead to identification of additional cases (Hellmich et al. 2017), but that the process was laborious. EMRs have also been used to monitor provider contact with infected persons by using information about which providers went into a particular patient room (Gannon 2020). While this is very interesting and valuable in the context of in-hospital infection prevention, it does not immediately address new clusters in the community.

Automated contract tracing via phone applications shows promise for the identification of contacts. This technology relies on the fact that many people have smart phones that can host applications. These applications can then record if a person has come in contact with an infected individual. The application can then signal the individual and/or public health officials regarding the contact and trigger the traditional isolation and support of the contact. Not without its faults, first that many people in a given community would be required to both have a smart phone and the application installed on said smart phone. There are unresolved privacy questions including what information is shared(i.e. how much information is stored and retained by the application developers and what information is provided to health departments or provided to other third parties) (Cho, Ippolito, and Yu 2020).

Additionally, analysis of SARS-CoV-2 has shown that the virus typically spreads in so-called super-spreader events (Liu, Eggo, and Kucharski 2020; Endo et al. 2020). This means that one index case may generate many secondary cases, while the majority of cases stop at a single case (Lloyd-Smith et al. 2005). Stopping these super-spreader events is important to ending an epidemic and the spread of infection in a community. Studies have shown that if contact tracing can be used to test, trace, and isolate certain high risk communities stopping the spread of infection can be made more effective (Eames 2007). However, contact tracing is limited by several factors including staffing. Providing rapid identification of super-spreading events and the associated clusters can ameliorate lack of staffing by focusing resources on known linked cases.

## 2 Methods

Data from the Electronic Medical Record is processed into a local enterprise data warehouse (EDW). For SARS-CoV-2 the critical data collected were positive test results. These positive tests were recorded both for on-site rapid tests and external laboratory results. These positive test results were then paired with general patient information collected about the patients within the EMR as part of the normal intake process. These data include the patient’s reported address, their primary spoken language, and employer. Employment history was also supplemented with insurance information (e.g. plan numbers) if the patient had commercial insurance.

The addresses on file for those patients were then geocoded using an on-premises secure software. Prior to geocoding the addresses, a program was run to clean the addresses to increase the probability of a matching the address in the geocoding server (e.g. consistent abbreviations for streets, removal of apartment numbers, amongst other common misspellings). These geocoded addresses were then truncated to four decimal places in order to represent a 30 foot radius around the reported address. A similar approach was used for employment information with self-reported employment information being cleaned for consistency as well as compared against a list of locally developed employer alias (e.g. Proctor & Gamble = Proctor And Gamble) in order to account for inconsistencies in naming. Consistent cleaning of employers also allows the employers to be matched against the North American Industry Classification System provided by the United States Bureau of Labor Statistics. Aggregating employers within this framework provides additional insight as to the types of work where outbreaks are occurring (e.g. manufacturing versus service).

Using understanding of local demography, a select group of languages were generated. It is known that certain immigrant communities settled in well defined areas of the service region. Because of the close-knit nature of these communities and the relative small number of speakers of these languages, these languages were also used to examine for potential outbreaks.

In order to establish possible linked cases, the positive patients were grouped on each of the three main criteria, location, employer, and language. For those persons that shared a unique location, a “cluster” was formed. Once a cluster group was identified, the transmission chain was probabilistically linked using a Bayesian framework as described in T et al. (2014) and implemented in the R package outbreaker2 (Campbell et al. 2018). Literature reported values were used for the serial interval (Nishiura, Linton, and Akhmetzhanov 2020). Application of this process resulted in an estimate of the transmission chain for the cluster. The highest probability transmission chain was taken as the likely transmission chain. This process was repeated for employers, language groups, and persons in large congregate settings (e.g. migrant housing communities, trailer parks). If the time between cases was greater than a threshold (e.g. 30 days), then the transmission chain was truncated.

The derived cluster information was combined from the three different sources and paired with other demographic information like age, race, and gender. If an individual appeared in more than one contact network, the earliest known transmission chain was retained. This data was then combined into a line list, or list of all positive cases, and a contact list that contained the above information regarding who likely was the index case for each infection. All data cleaning and aggregation took place in the R Statistical Environment (R Core Team 2019). The Epicontacts package (Nagraj et al. 2017) was utilized in order to visualize the transmission chains under a variety of different views.

Please see the appendix for implementation details.

## 3 Results

Automated contact tracing has highlighted several emerging outbreaks in the target communities for the health network. By applying this procedure, cases are probabilistically linked and local health departments and internal infectious disease teams can be alerted automatically as positive cases appear in the EMR. This has allowed for rapid identification of emerging clusters, and internal decisions made about mobile testing locations and community outreach to the effected community.

## 4 Discussion

### 4.1 Limitations

Automatic contact tracing can never take the place of the actual contact tracing process performed by tracers. Use of the EMR makes many assumptions. Among these assumptions are that the data available in the EMR is correct. Much of the information about address, language, race, ethnicity, are all manually collected and transcribed into the EMR. Strong assumptions are also made regarding the geocoding of addresses. The geocoding as is may place two or more people within a particular apartment building, but this does not necessarily mean that transmission occurred between these two people. Establishing the exact transmission chain is not possible using the EMR information. For that reason, the estimated transmission chains trade a higher probability of error for the speed of identification of emerging clusters.

### 4.2 Next Steps

This methodology, while developed on SARS-CoV-2 and the associated COVID-19 outbreak could also be used for other infectious diseases within a given community. This approach could surface potential influenza outbreaks as well as other common infectious diseases (mumps, measles, tuberculosis, etc). Monitoring these infectious diseases and quickly surfacing links cases would allow the health system and the department of public health to work together to manage outbreaks and craft at the earliest possible stages of spread within a community. Use of genetic sequences could also be employed to further refine estimation of the transmission chains as discussed in T et al. (2014). This would provide a much higher degree of certainty regarding likely transmission, but would require that the genetic sequencing information be made available in an EMR.

## 5 Conclusion

Automatic contact tracing and cluster identification using the EMR provides a methodology to probabilistically identify transmission chains and likely emerging clusters of cases. During a pandemic when the burden of contact tracing falls on local departments, use of such tools could help them focus their limited time and resources on emerging outbreaks.

## Data Availability

Implementation of the proposed methodology is available on GitHub.

https://github.com/conedatascience/contacttracerpaper

https://github.com/conedatascience/autotracer

## Footnotes

1 Also of note is Mossong et al. (2008) where depending on the country and age band, the number of contacts per day could be between 9–18 contacts per day

